# Decoupling sleep and brain size in childhood: An investigation of genetic covariation in the ABCD study

**DOI:** 10.1101/2020.10.02.20204735

**Authors:** Leanna M. Hernandez, Minsoo Kim, Cristian Hernandez, Wesley Thompson, Chun Chieh Fan, Adriana Galván, Mirella Dapretto, Susan Y. Bookheimer, Andrew Fuligni, Michael Gandal

**Affiliations:** Department of Psychiatry and Biobehavioral Sciences, Semel Institute, David Geffen School of Medicine, University of California Los Angeles, 695 Charles E. Young Drive South, Los Angeles, CA 90095, USA; Program in Neurobehavioral Genetics, Semel Institute, David Geffen School of Medicine, University of California, Los Angeles, Los Angeles, CA 90095, USA; Department of Family Medicine and Public Health, University of California, San Diego, San Diego, CA 92093, USA; Center for Human Development, University of California, San Diego, San Diego, CA 92093, USA; Department of Psychology, University of California, Los Angeles, Los Angeles, CA 90095, USA; Ahmanson-Lovelace Brain Mapping Center, University of California, Los Angeles, Los Angeles, CA 90095, USA; Staglin IMHRO Center for Cognitive Neuroscience, University of California, Los Angeles, Los Angeles, CA 90024, USA; Department of Neurology, Center for Autism Research and Treatment, Semel Institute, David Geffen School of Medicine, University of California, Los Angeles, 695 Charles E. Young Drive South, Los Angeles, CA 90095, USA; Department of Human Genetics, David Geffen School of Medicine, University of California, Los Angeles, Los Angeles, CA 90095, USA

## Abstract

**Background:** Childhood sleep problems are common and among the most frequent and impairing comorbidities of childhood psychiatric disorders. However, little is known about the genetic architecture of childhood sleep and potential etiological links between sleep, brain morphology, and pediatric-onset psychiatric symptoms.

**Methods:** Using data from the Adolescent Brain and Cognitive Development Study (N_Phenotype_=4,428 for discovery/replication, N_Genetics_=4,728, age: 9-10), we assessed phenotypic relationships, heritability, and genetic correlation between childhood sleep disturbances (SDs: insomnia, arousal, breathing, somnolence, hyperhidrosis, sleep-wake transitions), brain size (surface area [SA], cortical thickness, volume), and dimensional psychopathology.

**Results:** SDs showed widespread positive associations with multiple domains of childhood psychopathology; however, only insomnia showed replicable associations with smaller brain SA. Among the SDs assessed, only insomnia showed significant SNP-based heritability (*h*^*2*^_SNP_=0.15, *p*<0.05), and showed substantial genetic correlations with externalizing symptoms and attention-deficit hyperactivity disorder (ADHD; *r*_*G*_’s>0.80, *p*’s<0.05), suggesting significant pleiotropy across these complex childhood traits. We find no evidence of genetic correlation between childhood insomnia and brain size. Polygenic risk scores (PRS) calculated from genome-wide association studies (GWAS) of adult insomnia and adult brain size did not predict childhood insomnia; instead, PRS trained using ADHD GWAS predicted decreased SA at baseline, as well as insomnia and externalizing symptoms longitudinally.

**Conclusions:** These findings demonstrate a distinct genetic architecture underlying childhood insomnia and brain size and indicate that childhood insomnia should be considered along the dimensional axis of externalizing traits. Uncovering shared and unique genetic risk across childhood traits may inform our understanding of the developmental origins of comorbid psychiatric disorders.

## Introduction

Sleep is a fundamental biological process that is critical for optimal physical and cognitive development.^1–5^ The structure of sleep and circadian rhythms undergo substantial shifts across the lifespan, especially during critical adolescent periods when many psychiatric symptoms emerge. Sleep disturbance is nearly universally observed in childhood and adult psychiatric disorders and often among the most impairing comorbidities.^6^ In adults, genome wide association studies (GWAS) have shown that sleep-related behaviors are heritable and polygenic, with more than 200 associated genetic loci, and show significant genetic correlations with adult psychiatric disorders including depression and sczhizoprenia.^7–10^ Genetic variants associated with adult sleep disorders are enriched for genes expressed in the brain and have implicated pathways involved in locomotor behavior, neurodevelopment, and synaptic transmission, among others,^7–10^ suggesting a neurogenetic basis for individual differences in sleep behavior. Yet, while the heritable polygenic contributions to adult sleep disturbance have been established, much less is known about the origins of sleep-related traits in childhood, a developmental period during which sleep disruptions are extremely common, affecting an estimated 20-30% of the general pediatric population.^11–13^ Identifying a genetic basis for childhood sleep disturbance – and potential shared genetic etiology with neurodevelopmental, psychiatric disorders – would provide a critical foundation for understanding fundamental aspects of developmental biology and for fostering potential clinical interventions for the millions of children who experience sleep disturbances.

Childhood and adolescence are characterized by age-related decreases in brain volume, surface area, and cortical thickness,^14,15^ coinciding with normative shifts in the timing of homeostatic and circadian rhythms,^16^ and the emergence of life-long chronic psychiatric disorders.^17^ Thus, biological interactions between sleep, brain structure, and mental health may be particularly important during childhood, when dynamic changes occur in all three domains. Indeed, recent neuroimaging research suggests that childhood psychiatric symptoms (attention-deficit/hyperactivity disorder [ADHD],^18^ depression^19^) predict future sleep disturbances and moderate the association between smaller brain size and sleep problems. However, few large-scale genetics studies have been conducted to identify a heritable basis for sleep disturbance in childhood or to elucidate potential genetic relationships between sleep disturbance and childhood mental health,^18–20^ leaving unanswered how polygenic risk for sleep, brain, and psychiatric traits interact over the course of development. By contrast, there has been robust and well-powered genomics research in adults indicating significant genetic pleiotropy between adult insomnia, sleep duration, neuropsychiatric disorders (e.g., depression, anxiety, ADHD, and schizophrenia^7,8,10^), and brain surface area,^21^ indicating that genetic risk for these traits in adulthood is in-part shared. Parallel analyses of the shared genetic architecture between childhood sleep psychiatric symptoms, and brain structure have not been performed and may provide novel insights into the developmental origins of comorbid psychiatric/brain traits.

To address the these questions, we perform a comprehensive examination of the phenotypic and genetic relationship between sleep, mental health, and brain structure in youth who participated in baseline and follow-up assessments of the Adolescent Brain and Cognitive Development (ABCD) study.^22^ Using parent-report data of children’s sleep behavior (i.e., Sleep Disturbance Scale for Children^23^) and behavioral/psychiatric symptoms (i.e., Child Behavior Checklist^24^), we determine the extent to which individual variability across 6 dimensions of sleep disturbance (i.e., insomnia, arousal, breathing, somnolence, hyperhidrosis, sleep-wake transitions) predict individual variability in 20 subscales of psychiatric symptomatology, and magnetic resonance imaging (MRI) measures of brain volume, surface area, and cortical thickness at 9-10-years of age. To estimate the contribution of common genetic factors, we calculate the heritability of – and genetic correlations between – childhood sleep, psychiatric symptoms, and global brain size. Finally, we examine the extent to which polygenic risk scores (PRS) for sleep, brain, and psychiatric traits predict brain and behavioral phenotypes in children at 9-10-years of age and psychiatric symptoms 1-year later.

## Methods and Materials

### Subjects

Participants were youth who completed the baseline and 1-year follow-up assessment of the ABCD study,^22^ an ongoing multi-site longitudinal investigation of brain development in the United States. Subjects were recruited from schools within geographical areas that approximate the sociodemographic diversity of the national population.^25^ The current analysis was completed using de-identified structural neuroimaging, genetic, demographic and behavioral data obtained from the ABCD 2.0.1 National Data Archive (NDA) data release (NDAR DOI: 10.15154/1504041).

### Sleep Disturbance Scale for Children (SDSC)

The SDSC^23^ was used to generate dimensional indices of sleep problems. The SDSC is a parent-report questionnaire designed to identify the presence of sleep disturbances over the past 6 months in children with/without clinically significant sleep disorders. Individual item-level scores on the SDSC are grouped into 6 sleep-disorder subscales: Disorders of Initiating and Maintaining Sleep (DIMS), Sleep Breathing Disorders (SBD), Disorders of Arousal (DA), Sleep-Wake Transition Disorders (SWTD), Disorders of Excessive Somnolence (DOES), and Sleep Hyperhidrosis (SHY). The DIMS subscale encompasses a single question relating to sleep duration, as well as several questions indexing sleep quality (e.g., insomnia-related symptoms); we focused our analysis on DIMS questions indexing insomnia-related symptoms (Supplementary Materials). For all SDSC sleep measures, higher scores are indicative of more severe sleep problems.

### Child Behavior Checklist (CBCL)

The CBCL^24^ is a widely used parent-report questionnaire indexing child behavior across a number of psychiatric domains. Here, we assess the relationship between sleep and t-scores on 14 CBCL subscales, as well as 6 DSM-oriented subscales.

### NIH Toolbox Cognition Battery

Cognitive function was assessed using the NIH Toolbox® Cognition Battery,^26^ which was administered to children on an iPad. Summary scores indexing Cognitive Function Composite Score, Fluid Cognition Composite Score, and Crystalized Cognition Composite Scores were obtained for each child.

### Structural Magnetic Resonance Imaging (sMRI)

Structural MRI (sMRI) data were collected by individual sites affiliated with the ABCD consortium on either a Siemens Prisma, Phillips, or GE 750 3T scanner (see Supplementary Materials for scan parameters).^27^ Morphometric measurements of cortical thickness (CT), surface area (SA), and subcortical volume (VOL) were calculated in FreeSurfer using the Desikan parcellation atlas.^28^ As recommended in the ABCD NDA 2.0.1 Release Notes for Imaging Instruments, exclusionary criteria included subjects with MRI findings that were considered for clinical referral (N=402), subjects with poor quality T1 images (as described above; N=11), and subjects for whom the Freesurfer parcellation performed poorly (N=391).

### Phenotypic Associations

To demonstrate the robustness of the reported phenotypic associations, the sample was split into discovery and replication cohorts. One child per twin/triplet was included to ensure the statistical independence of the data in each of the cohorts; siblings were randomly assigned. The final sample for phenotypic association analyses consisted of 8,856 subjects, which were split into discovery (N=4,428) and replication (N=4,428) cohorts using the ISLR package in R (demographics provided in Supplementary Table 1). Results were deemed to be significant and to replicate if they survived FDR correction (*q*<0.05) in the discovery cohort and demonstrated a *p*<0.05 in the replication cohort.

Statistical analyses were performed in R v4.0.2. Linear mixed-effects models were run using the lme4 package to test the relationship between SDSC subscales and sMRI indices. Dependent variables included CT and SA (34 regions per hemisphere), as well as 39 measures of subcortical VOL. Independent variables were sleep-related phenotypes (i.e., insomnia, SBD, DA, SWTD, DOES, and SHY). Age, socioeconomic status (SES; computed as the average of parental education and family income), sex, and parent-reported ethnicity were specified as fixed effects; family ID (to control for sibling relatedness) and MRI scanner were modeled as random effects. Analyses were performed with/without controlling for global sMRI measures, by including either whole brain VOL, mean SA, or mean CT as covariates. The relationship between CBCL and SDSC subscales were assessed using a series of log-linked gamma general linear models (GLM) and identical covariates.

### Genotyping

DNA was extracted from saliva samples and genotype data for 733,293 single nucleotide polymorphisms (SNPs) was generated using the Affymetrix NIDA Smokescreen Array. Sample preparation and genotyping was performed at the Rutgers University Cell and DNA Repository (RUCDR; see Supplementary Materials). The ABCD DAIC applied quality control (QC) filters based on call signals and call rates and removed individuals who had a missing rate of greater than 20%, or for whom there may have been sample contamination. The NDA download thus consisted of genotype data for 10,627 subjects across 517,724 unique SNP markers.

### SNP-Based Heritability and Genetic Correlations

Ancestry was determined by merging ABCD data with data from the 1000 Genomes Project^29^ followed by principal component analysis of the merged dataset and application of a k-nearest neighbors classification algorithm to the first four ancestry principal components. Subsequent analyses were restricted to subjects of European ancestry to control for population stratification. PLINK v1.90^30^ was used to prune SNPs for linkage disequilibrium (LD), minor allele frequency <1%, missingness per individual >10%, missingness per marker >10%, and Hardy-Weinberg equilibrium (*p*<10^−6^). The genetic relationship matrix (GRM) was computed from autosomal chromosomes using GCTA v1.93.^31^ One subject from each pair of individuals with an estimated relatedness greater than 0.05 was removed from further analyses, leaving 4,728 unrelated European subjects and 157,123 genotyped SNPs that passed QC. Ancestry principal components (PCs) for the European cohort were calculated in PLINK v1.90.

Heritability was estimated using GCTA’s restricted maximum likelihood (REML). Due to non-normality, SDSC subscales were binarized; subjects scoring in the top 25% of for each sleep subscale were defined as experiencing sleep problems (i.e., cases) and the remainder served as a control group. A rank-based inverse normal transformation was applied to continuous phenotypes (i.e., cognition and neuroimaging variables) to correct for deviations from normality. For heritability estimates of behavioral and cognitive measures, age, SES, sex, 10 ancestry PCs, and genotyping batch were included as covariates. Heritability estimates for neuroimaging phenotypes indexing brain size used identical covariates in addition to MRI scanner. Bivariate genetic correlations were calculated in GCTA^32^ with identical covariates.

### Polygenic Risk Score Profiling

To assess the effects of subject-level genetic liability, polygenic risk scores (PRS) were computed for European subjects using PRS-CS^33^ with summary statistics from GWAS of insomnia,^7^ ADHD,^34^ and brain surface area.^21^ Genotype data was imputed using the Michigan Imputation Server^35^ using the TOPMed reference panel with Eagle v2.3 phased output and mixed ancestry for QC. Individuals with >10% missing genotypes, and SNPs with >10% missingness rate, minor allele frequency <5%, or out of Hardy-Weinberg equilibrium (*p*<10^−6^) and SNPs in the major histocompatibility complex were removed were removed, leaving 4,534,127 genetic variants. A relatedness cutoff of 0.05 was applied, leaving 3,386 subjects at baseline and 1,669 at follow-up. PRS were generated using the European LD reference and a global shrinkage parameter of 1e-2. The extent to which youth’s PRS predicted the binary insomnia phenotype was assessed using logistic regression controlling for age, SES, sex, and 10 ancestry PCs. Predictions of baseline and follow-up CBCL subscales were assessed using log-linked gamma GLM using identical covariates. Linear mixed-effects models were used to assess the relationship between PRS and brain structure, using an additional covariate of MRI scanner. All regressions using follow-up data were controlled for sleep or psychiatric symptoms at the first timepoint.

## Results

### Widespread Associations between Child Sleep Disturbance and Psychiatric Symptoms

Phenotypic associations between sleep and psychiatric symptoms were tested in large multi-ethnic discovery and replication cohorts (N’s=4,428). Regression analyses revealed robust and replicable positive associations; greater frequency of any sleep disturbance predicted higher levels of psychiatric symptomatology across all 20 CBCL subscales (*β’s*=0.01-0.04, discovery *q*<0.05, replication *p*<0.05; Figure 1; Supplementary Table 2-7), demonstrating broad associations between childhood SDs and behavioral/emotional problems in children. In addition, a small, but significant negative relationship was observed between insomnia and fluid cognition (*β*=-0.003, discovery *q*<0.05, replication *p*<0.05; Figure 1; Supplementary Table 2).

**Figure 1.**
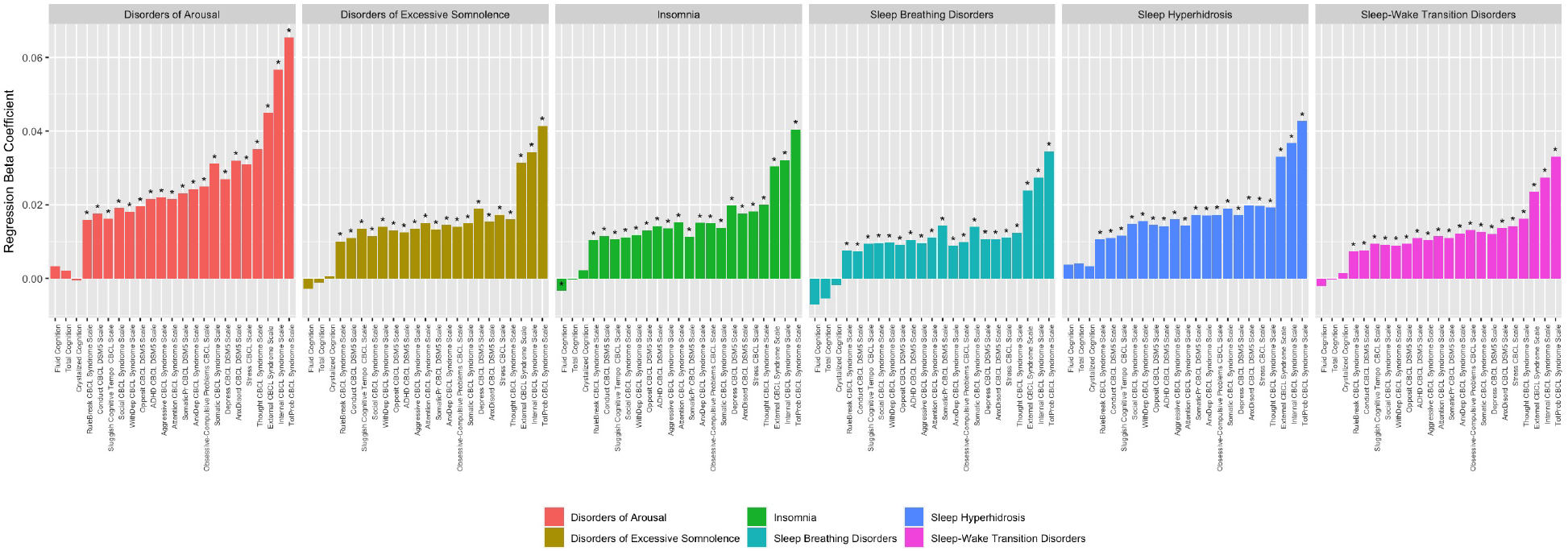
Broad phenotypic associations between childhood sleep and psychiatric symptoms. Sleep disturbances show replicable associations (discovery [N=4,428], replication [N=4,428]) with Child Behavior Checklist (CBCL) subscales of behavioral/emotional problems and NIH Toolbox fluid cognition. Variables showing a significant association with sleep subscales in the discovery cohort after FDR correction (*q*<0.05) and demonstrating a nominally significant association in the replication cohort (*p*<0.05) are denoted by an asterisk (*). Data shown reflects associations in the discovery cohort. For statistical associations in the replication cohort, see Supplementary Materials.

### Childhood Insomnia is Uniquely Associated with Decreased Brain SA, VOL

We next examined the relationship between childhood SDs and global measures of brain structure (mean CT, mean SA, and whole brain VOL) at baseline. More frequent insomnia showed replicable associations with lower SA and smaller VOL (discovery *q*<0.05, replication *p*<0.05; Figure 2A; Supplementary Table 8). No other SD showed replicable associations with global brain structure, nor were any measures associated with cortical thickness. As such, childhood insomnia was uniquely associated with global measures of childhood brain structure.

**Figure 2.**
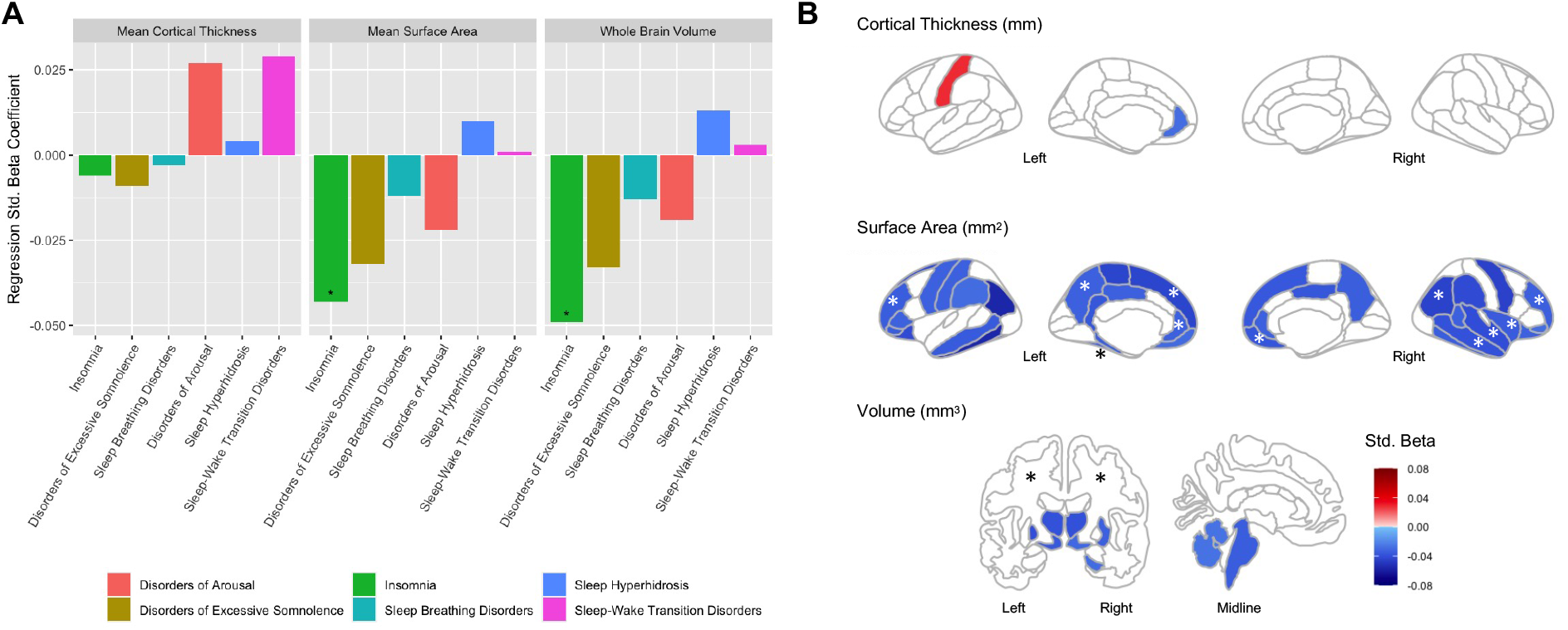
More frequent insomnia symptoms is associated with smaller brain size in 9–10-year-old youth. **A)** Associations between sleep disturbances and global measures of mean cortical thickness, mean surface area, and whole brain volume. Variables showing a significant association with sleep subscales in the discovery cohort (N=4,428) after FDR correction (*q*<0.05) and demonstrating a nominally significant association in the replication cohort (N=4,428; *p*<0.05) are denoted by an asterisk (*). **B)** Brain regions demonstrating a significant association with insomnia in the discovery cohort (*q*<0.05) are shown in color; regions also demonstrating a significant association in the replication cohort (*p*<0.05) are denoted by an asterisk (*). In both panels, plotted data reflects associations in the discovery cohort. For statistical associations in the replication cohort, see Supplementary Materials.

To test for regional associations between SDSC sleep domains and brain structure, linear mixed-effects models were run with regional-measures of CT, SA, and VOL as dependent variables and each sleep-related phenotype as an independent variable. Greater frequency of insomnia symptoms was related to reduced SA in frontal, temporal and parietal regions, as well as smaller VOL of cerebral white matter (Figure 2B; Supplementary Tables 9-10). Regional findings were no longer significant when controlling for global measures, indicating that the association between insomnia and whole-brain SA and VOL is an important contributor to the observed region-level effects (Supplementary Tables 11-12). No other SDSC subscales demonstrated replicable effects (Supplementary Tables 13-29).

### Childhood Insomnia Exhibits Significant SNP-based Heritability

We next sought to determine the contribution of common genetic variation to childhood sleep, psychiatric symptoms, and brain structure. Among childhood sleep-related phenotypes, significant SNP-based heritability was observed for insomnia only (*h*^*2*^_SNP_=0.15, *p*<0.05; Figure 3A, Supplementary Table 30). Significant SNP-based heritability was also observed for CBCL subscales indexing ADHD, attention problems, somatic problems, externalizing, oppositional defiant disorder, and total problems (*h*^*2*^_SNP_=0.15-0.22, *p*’s<0.05), NIH Toolbox measures of fluid, crystalized, and fluid cognition (*h*^*2*^_SNP_=0.29-0.36, *p*’s<0.001), and neuroimaging measures of whole brain volume, mean surface area, and mean cortical thickness (*h*^*2*^_SNP_=0.27-0.37, *p*’s<0.01).

**Figure 3.**
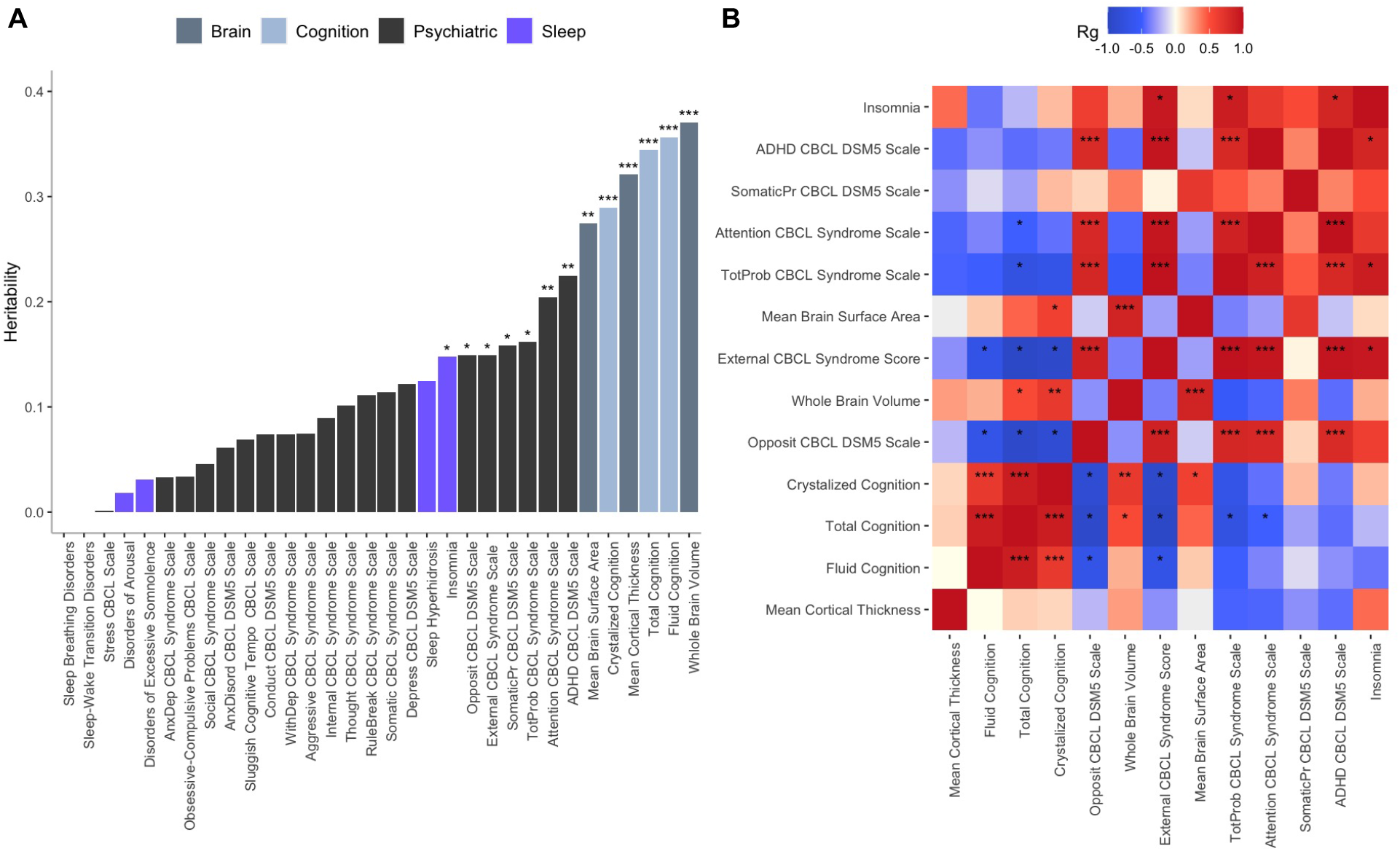
Genetic analyses indicate that childhood insomnia is heritable and demonstrates significant genetic correlation with ADHD and externalizing symptoms. **A)** SNP-based heritability of childhood sleep-related behaviors, psychiatric symptoms, and brain size (N=4,728 [European ancestry]). Among sleep-related traits, only insomnia demonstrates significant SNP-based heritability (*h*^*2*^_SNP_=0.15, *p*<0.05). **B)** Genetic correlations between pairs of traits that displayed significant SNP-heritability. Childhood insomnia shows significant genetic pleiotropy with attention deficit hyperactivity disorder (ADHD), externalizing, and total problems subscales of the Child Behavior Checklist (CBCL; *rG*’s>0.84, *p*’s<0.05; N=4,728 [European ancestry]). \**p*<0.05, \*\**p*<0.01, \*\*\**p*<0.001.

### Genetic Overlap between Childhood Insomnia, ADHD, and Externalizing Behaviors

Given the observed heritability of childhood insomnia, and strong genetic association between adult insomnia and brain structure^21^ we next investigated potential shared genetic influences on psychiatric symptoms and brain structure in childhood. Pairwise genetic correlations were performed between traits that demonstrated significant SNP-heritability. Significant positive genetic correlations were observed between insomnia and CBCL total problems, externalizing, and ADHD (*r*_*G*_’s>0.84; *p*’s<0.05; Figure 3B; Supplementary Table 30). For neuroimaging traits, significant positive correlations were observed between whole-brain VOL and mean SA, crystalized, and total cognition, and between mean SA and crystalized cognition (*r*_*G*_’s>0.53, *p*’s<0.05; Supplementary Table 31). Notably, however, we did not detect significant genetic correlations between childhood insomnia and brain CT, SA, or VOL. These results indicate a shared genetic basis among childhood insomnia, ADHD, and externalizing behaviors that does not contribute to decreased brain size.

### Distinct Genetic Contributions to Childhood versus Adult Insomnia

The previous data identify a heritable basis for childhood insomnia, exhibiting genetic and phenotypic overlap with ADHD/externalizing symptoms. To replicate and extend these associations, we calculated polygenic risk scores (PRS) using the largest available GWAS summary statistics of insomnia (in adults) and ADHD. Surprisingly, PRS trained using weights derived from *adult* insomnia GWAS did not predict the frequency of *childhood* insomnia symptoms at 9-10-years of age (*β*_*Std*._=0.006, *p*=N.S.; Figure 4A), frequency of insomnia symptoms at follow-up 1-year later (*β*_*Std*._=0.005, *p*=N.S.), or the binary insomnia phenotype at either timepoint (Baseline *β*_Std._=0.07, Follow-up *β*_Std._=0.04, *p*’s=N.S.; Figure 4B). The insomnia PRS did, however, predict baseline ADHD symptoms (*β*_*Std*._*=*0.006, *p*<0.01; Figure 4B), indicating that the GWAS was adequately powered to detect phenotypic associations. Mirroring our observed genetic correlations, we found that higher ADHD PRS was associated with the binary insomnia phenotype at both baseline (*β*_*Std*._=0.11, *p*<0.05) and 1-year follow-up timepoints (*β*_*Std*_.=0.15, *p*<0.05; Figure 4B). As a positive control, ADHD PRS also significantly predicted baseline and follow-up externalizing (Baseline *β*_Std._=0.02, Follow-up *β*_Std._=0.01, *p*’s<0.05) and ADHD symptoms (Baseline *β*_Std._=0.01, Follow-up *β*_Std._=0.01, *p*’s<0.01; Figure 4B). Together, these results demonstrate that the common genetic variants contributing to the heritability of insomnia symptoms in adults are largely distinct from those in children.

**Figure 4.**
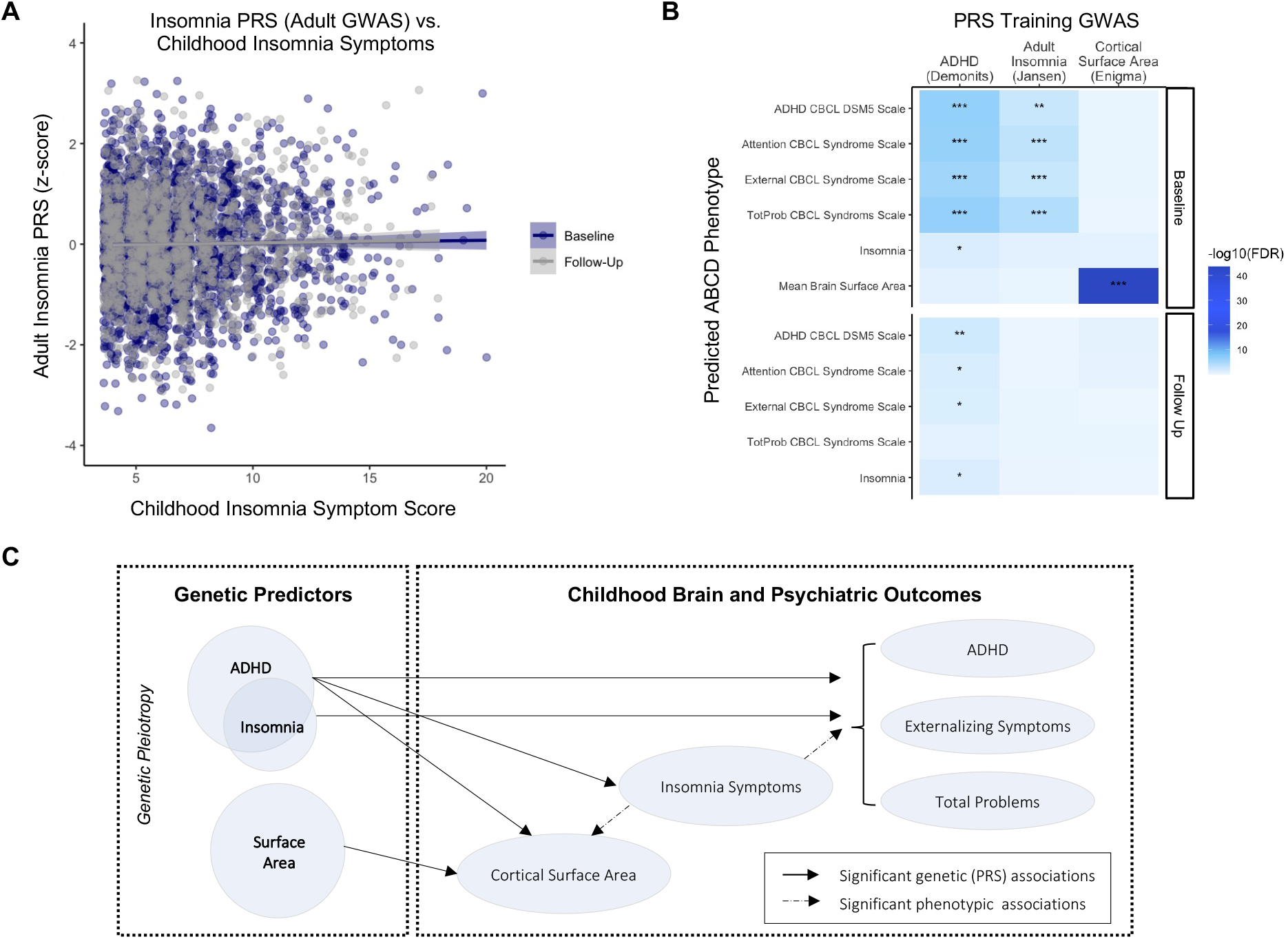
Polygenic risk for ADHD – but not insomnia – contributes to smaller brain size, insomnia, and externalizing behaviors in childhood. **A)** Polygenic risk scores (PRS) trained using weights from GWAS of insomnia (Jansen et al., 2019) in adulthood do not predict insomnia symptoms in 9-10-year-old children (baseline) or at follow-up 1-year later (*p*’s=N.S.). **B)** PRS trained using GWAS of ADHD (Demontis et al., 2019) predict not only externalizing-related behaviors at baseline and follow-up, but also predict insomnia at both time points. **C)** Proposed model of associations between genetic risk, sleep, and mental health outcomes. The pleiotropic effects of shared genetic risk between ADHD and insomnia drive observations of smaller brain surface area, more frequent insomnia symptoms, and childhood externalizing symptoms. \**p*<0.05, \*\**p*<0.01, \*\*\**p*<0.001.

### Insomnia-Brain Structure Associations are Mediated by ADHD PRS

Finally, we sought to better understand the observed, replicated phenotypic associations between insomnia and decreased brain VOL/SA. Here, we generated PRS to predict the genetic component of brain SA, using the largest available GWAS from ENIGMA.^21^ As a positive control, SA PRS very strongly predicted brain SA in the ABCD cohort (*β*_*std*_=0.23, *p*<0.001; Figure 4B). In contrast, SA PRS was not associated with childhood insomnia symptoms (*β*_*Std*_=0.08, *p*=N.S.), affirming our null genetic correlation results. Previous work has identified a negative genetic association between ADHD and intracranial volume.^36^ To determine whether genetic overlap with ADHD could account for our observed insomnia-brain structure associations, we performed a series of follow-up analyses in brain regions where we found replicable phenotypic associations (i.e., regions shown in Figure 2B). Notably, ADHD PRS significantly predicted lower SA in three regions — left superior frontal gyrus (*β*_*Std*._=-0.04, *p*<0.05), left rostral middle frontal gyrus (*β*_*Std*._=-0.04, *p*<0.05), and right insula (*β*_*Std*._=-0.03, *p*<0.05). Additionally, we found that the association between insomnia and brain SA disappeared when conditioning on ADHD. Together, these findings support a dissociable pleiotropic model in which genetic risk for ADHD contributes separately to both smaller brain SA and symptoms of insomnia in childhood, such that insomnia symptoms are not directly mediated by brain structure (Figure 4C). In support of this model, we further observed that ADHD PRS continues to predict the binary insomnia phenotype at baseline and follow-up timepoints even when controlling for brain structure (i.e., mean cortical SA; Baseline *β*_*Std*._=0.11, Follow-up *β*_*Std*._=0.15, *p*’s< 0.05) and its potential genetic drivers (i.e., SA PRS; Baseline *β*_*Std*._=0.11, *β*_*Std*._=0.15, *p*’s<0.05). Altogether, these findings greatly refine our understanding of the genetic architecture of childhood sleep disturbance, as well as its relationship to adult sleep disturbance, childhood brain structure, and developmental psychopathology.

## Discussion

Leveraging longitudinal data from a population-based, demographically diverse sample of youth, the current study examined phenotypic relationships, heritability, and genetic correlations between childhood sleep disturbances, brain size, and dimensional measures of psychopathology. Among the six domains of childhood sleep disturbances examined (i.e., insomnia, arousal, breathing, somnolence, hyperhidrosis, sleep-wake transitions), only childhood insomnia showed replicable associations with brain size and was heritable (*h*^*2*^_*SNP*_ 0.15). We demonstrate positive genetic correlations between insomnia and ADHD/externalizing behaviors, indicating shared genetic etiology (i.e., pleiotropy) across these complex childhood traits. At the phenotypic level, the frequency with which youth experienced insomnia symptoms was broadly associated with greater parent-reported psychiatric symptomatology across 20 subscales of mental health and was negatively associated with structural neuroimaging measures of whole-brain volume, mean cortical surface area, and regional surface area in frontal, parietal and temporal cortices. Notably, despite these phenotypic associations, we did not observe genetic correlations between childhood insomnia and brain size, suggesting distinct genetic mechanisms. Further, PRS trained using weights from adult GWAS of brain size did not predict childhood insomnia symptoms. Rather, insomnia could be predicted by PRS for ADHD, even when controlling for brain structure. Together, our results show that childhood insomnia is not mediated by reduced brain size, which rather appears to reflect the pleiotropic effects of shared ADHD genetic risk. These findings highlight a model of shared polygenic mechanisms underlying sleep quality and externalizing traits in childhood and suggest that childhood insomnia should be classified along the dimensional axis of ADHD/externalizing disorders.

One mechanism through which sleep may affect trajectories of brain development is through alterations of normative developmental processes, including synaptic pruning. Age-related decreases in cortical thickness, surface area, and volume occur throughout childhood and adolescence,^14,15^ which are thought to be driven by ongoing synaptic pruning^37^ and consequent changes to brain area and gyrification.^38^ In mice, chronic sleep deprivation has been associated with upregulation of astrocytic phagocytosis and microglial activation, resulting in enhanced synaptic degradation.^39^ Thus, long-term disturbances in sleep may promote prolonged microglial activation, making the brain more susceptible to other genetic and environmental insults.^39^ Indeed, we found that poorer sleep – across multiple dimensions of sleep behavior – is consistently associated with higher levels of parent-endorsed psychiatric symptoms. Our multi-trait sleep analysis extends the finding of recent population-representative pediatric sleep studies, which have either narrowly focused on sleep duration (i.e., hours of sleep per night)^19,20^ or on a limited number of psychiatric traits,^18^ and suggest that behavioral interventions aimed at improving the full range of sleep issues experienced by youth may improve mental health across the clinical spectrum.

Neural regulation of sleep and wakefulness is governed by reciprocal communication between the brain stem, subcortical regions, and cortex.^40^ In line with previous research findings of reduced grey matter volume in sleep disturbed children^41^ and adults^42^, we find that childhood insomnia is associated with reduced surface area in the frontal cortex (among other cortical regions), a brain area encompassing the basal forebrain, which animal studies have shown contain GABAergic neural populations critical for wakefulness.^43,44^ Recent work has further demonstrated the relationship between brain size and sleep is mediated by psychiatric symptoms.^18,19^ Here, we refine this model through the addition of genetics, which provides a causal directional anchor. We find that it is ADHD/externalizing psychiatric symptoms and their genetic risk (in particular) that mediate the relationship between brain size and sleep.

The SNP-based heritability of childhood insomnia was moderate (*h*^*2*^_SNP_=0.15) in the current pediatric sample, similar to estimates previously reported in adults (*h*^*2*^_SNP_=0.07-0.17).^7,45^ We also observed similar estimates of heritability for childhood cognition (*h*^*2*^_SNP_=0.29-0.36) and brain size (cortical thickness, cortical surface area, brain volume; *h*^*2*^_SNP_=0.27-0.37) as those documented in large-scale genomics studies of adult educational attainment (*h*^*2*^_SNP_=0.22)^46^ and brain structure (*h*^*2*^_SNP_∼0.26-0.34).^21,47^ In adults, common genetic variants affecting sleep, brain size, and mental health show high genetic correlations, suggestive of substantial pleiotropy.^7,21,45^ To test whether the heritable polygenic contributions to childhood sleep quality also affect childhood brain size, we generated bivariate genetic correlations between sleep-brain-psychiatric traits in our 9-10-year-old sample. Surprisingly, and in contrast to the adult literature, genetic correlations between childhood insomnia and brain size (cortical thickness, surface area, volume) did not support the existence of genetic pleiotropy.

To further validate these null results, we used SNP-weights derived from large-scale GWAS of adult insomnia and cortical surface area to generate individual-level PRS’s in the ABCD sample. Mirroring our null genetic correlations, PRS for brain surface area did not predict childhood insomnia. Instead PRS for ADHD was a significant predictor of *both* insomnia (at both 9-10-years of age and at the 1-year follow-up) and surface area in the frontal cortex and insula, which is consistent with recent studies showing genetic associations between ADHD, narcolepsy,^48^ and sleep disturbances more broadly.^49^ Our findings of positive genetic correlations between childhood sleep disturbance and ADHD are also in agreement with previous research documenting behavioral comorbidity^50,51^ and overlapping affected neural systems.^18^ We hypothesize that the genetic relationship between ADHD and childhood insomnia induces a phenotypic correlation between childhood insomnia and brain size; that is to say, as ADHD was pleiotropically associated with reduced brain surface area *and* insomnia symptoms, we hypothesize that this genetic relationship induces a phenotypic correlation between insomnia and brain size. Altogether, these data indicate that despite similar heritability estimates, childhood and adult insomnia show distinct genetic correlations with other brain and behavioral traits and suggest that different genetic mechanisms may underlie the relationship between childhood sleep/neurodevelopmental *vs*. adult sleep/adult-onset psychiatric disorders. Ultimately, however, multiple waves of longitudinal data will be required to fully elucidate the temporal relationship between sleep, brain size, and cognitive/psychiatric symptoms throughout the lifespan.

In sum, using publicly available data from a socio-demographically diverse sample of youth, we sought to elucidate the phenotypic and genetic associations between sleep, mental health, and brain structure. Overall, these data indicate that childhood insomnia is genetically rooted in polygenic risk for ADHD and that the link between genetic risk for sleep disturbance, sleep behavior, and psychiatric symptoms may change across the lifespan, underscoring the importance of further elucidating the relationship between sleep-related genes, brain, and behavior in developmental populations. As sleep is crucial for optimal cognition and mental health, a better understanding of the genetic and neural underpinnings of sleep disturbances in children is essential to further our understanding the relationship between sleep and brain development, informing public policy surrounding guidelines for optimal sleep during childhood, and designing effective interventions aimed at improving cognitive and psychiatric outcomes in youth who experience sleep difficulties.

## Data Availability

Data is available at the NIMH Data Archive (NDA)

https://nda.nih.gov/

