## Supplementary Materials for "Decoupling sleep and brain size in childhood: An investigation of genetic covariation in the ABCD study"

**Author Disclosures:** All authors report no biomedical financial interests or potential conflicts of interests.

**Corresponding author:** Michael Gandal, MD, Ph,D, Assistant Professor, Neuroscience Research Building, 635 Charles E Young Dr S, Los Angeles, CA 90095, University of California, Los Angeles,; Leanna Hernandez, PhD, Postdoctoral Fellow, Neuroscience Research Building, 635 Charles E Young Dr S, Los Angeles, CA 90095, University of California, Los Angeles,

### Methods

#### *Subject characteristics*

Demographic data obtained from the NDA included age (months), sex (male/female), parental education, family income, family ID/relationship status (i.e., an indication of subjects who are siblings), self-reported race/ethnicity, and MRI scanner serial number (Supplementary Table 1).

#### *Sleep Disturbance Scale for Children*

An insomnia score was computed for each subject by adding scores across 1) a single question that asked "How long after going to bed does your child usually fall asleep?," for which parents could choose one of five choices ranging from "less than 15 minutes" to "more than 60 minutes", and 2) three questions for which parents made Likert-style choices ranging from 1 (never) to 5 (always/daily): "The child has difficulty getting to sleep at night"; "The child wakes up more than twice per night"; "After waking up at night, the child has difficulty to fall asleep again." Insomnia scores thus ranged from 4-20 and did not include total sleep duration.

#### *Structural magnetic resonance imaging*

3D T1-weighted images had the following scan parameters: 3D T1-weighted images had the following scan parameters: Siemens - matrix size 256x256, 176 slices, FOV 256x256, resolution 1mmx1mmx1mm, TR 2500ms, TE 2.88ms, flip angle 8°, total scan time 7:12; Prisma - matrix size 256x256, 225 slices, FOV 256x240, resolution 1mmx1mmx1mm, TR 6.31ms, TE 2.9ms, flip angle 8°, total scan time 5:38; GE - matrix size 256x256, 208 slices, FOV 256x256, resolution 1mmx1mmx1mm, TR 2500ms, TE 2ms, flip angle 8°, total scan time 6:09.

All structural neuroimaging data were processed by the ABCD Data Analysis and Informatics Core (DAIC) and extensive documentation of image acquisition[1] and processing[2] methods is provided elsewhere. Briefly, quality control of sMRI images included a manual review of images for artifacts including wrap-around, missing brain due to improper slice prescription, signal drop out due to magnetic susceptibility artifacts, and motion. Tabulated ABCD sMRI data included a binary QC code for each participant (0=reject, 1=accept), which was used to remove subjects with poor quality sMRI data. Reconstruction of the cortical surface and brain segmentation was performed in FreeSurfer v5.3 (http://surfer.nmr.mgh.harvard.edu/). Morphometric measurements of cortical thickness (CT), surface area (SA), and subcortical volume (VOL) were calculated in FreeSurfer using the Desikan parcellation atlas.[3] Quality control of FreeSurfer data was performed by trained reviewers based on the presence of motion, intensity inhomogeneity, white matter underestimation, pila overestimation, and the presence of magnetic susceptibility artifacts. Tabulated sMRI data included a binary FreeSurfer QC score for each participant (0=reject, 1=accept), which was used to remove subjects with poor quality data.
